# Dynamic Prediction to Predict Virologic Failure in Antiretroviral Treated People Living with HIV: Cohort Analysis of Routine Electronic Health Records Data in the Western Cape, South Africa

**DOI:** 10.64898/2026.09.08.26362496

**Authors:** Frissiano Honwana, Elton Mukonda, Freedom Gumedze, Nei-yuan Hsiao, Landon Myer, Maia Lesosky

## Abstract

**Objectives:** Virologic failure (VF) remains a major challenge in HIV care, particularly in resource-limited settings where timely identification is critical. Dynamic prediction models can provide continuously updated, individual-level risk estimates using longitudinal biomarker data. However, their applicability to routinely collected electronic health record (EHR) data remains poorly understood. We developed and internally validated a dynamic prediction model for VF using routine EHR data from people with HIV (PWH) receiving antiretroviral therapy in South Africa.

**Methods:** A joint model (JM) linking a linear mixed-effects model for longitudinal HIV viral load (HIVVL) to a Cox model for time to first VF, adjusted for baseline age group and sex at birth, was developed using routine National Health Laboratory Service data from 122,425 PWH in the Western Cape, South Africa (SA) (January 2008-September 2018), randomly split into development (n = 91,818; 75%) and validation (n = 30,607; 25%) cohorts. Predictive performance was evaluated using time-dependent area under the receiver operating characteristic curve (AUC) and Brier scores at 12 and 24 months.

**Results:** Overall, 12,547 PWH (10.2%) experienced VF. The current value-parameterized model was used and showed moderate discrimination (AUC: 0.69 and 0.73) and good calibration (Brier scores: 0.033 and 0.051) at 12 and 24 months. Dynamic predictions updated meaningfully at each visit, demonstrating potential to flag individuals at elevated VF risk for timely clinical intervention.

**Conclusions:** Dynamic prediction models hold genuine promises to support personalized, timely clinical decision-making for PWH. However, EHR data with sparse measurements, missing clinical covariates, and irregular follow-up limit discriminative performance despite adequate calibration, demonstrating that data quality and covariate availability are at least as consequential as modelling strategy in real-world prediction settings.

## 1. Introduction

HIV remains a major public health burden in sub-Saharan Africa (SSA), accounting for 71% of the global burden of HIV infection despite the availability of effective antiretroviral therapy (ART) [1]. South Africa (SA) bears the largest share, contributing 23% of SSA incidence [1], with 77% of 7.7 million people with HIV (PWH) in the country estimated to be on ART in 2023 [2]. Lifelong treatment requires sustained adherence to prevent drug resistance [3, 4], and HIV viral load is the preferred biomarker for monitoring treatment response [4]. Virologically non-suppressed individuals (HIV viral load ≥1000 copies/mL) risk progressing to virologic failure (VF), an outcome associated with worse quality of life and fewer treatment options when not detected early [5]. Although viral load testing has historically been constrained by cost and logistical complexity in resource-limited settings [6, 7], SA’s annual test volume grew from 1.96 million in 2013 to 6.27 million in 2022, consistent with national guidelines recommending at least annual testing [8, 9].

The expanding volume of longitudinal viral load population test results creates a meaningful opportunity. Rather than relying on a single baseline measurement, repeated observations over time can provide richer information about individual disease trajectories and support more accurate, and timely prognosis prediction. Most existing prediction models for HIV-related outcomes use population-level or subgroup averages and focus predominantly on outcomes such as immune reconstitution inflammatory syndrome [10], mortality [11, 12], CD4/CD8 ratio recovery [13], with limited attention to VF. The few VF-specific models that exist, from Ethiopia [14] and SA [15] rely solely on baseline biomarker values, which cannot capture the dynamic nature of HIV on ART-treated PWH.

Dynamic prediction models can address this limitation directly. Built on observed longitudinal biomarker trajectories, they permit prognosis prediction that updates continuously as new measurements accumulate [16, 17]. Joint models for longitudinal and time-to-event data provide a principled statistical framework for this purpose, linking a mixed-effects model for the individual biomarker trajectory to a Cox proportional hazards model for the event outcome [16–19]. Applications have spanned CD4-based predictions in HIV [16, 20], prostate-specific antigen trajectories in prostate cancer [21, 22], and a range of conditions in resource-rich settings [23–25]. These models are usually applied to research cohort data collected under controlled conditions with systematic follow-up and comprehensive covariate coverage.

Whether the performance advantages attributed to dynamic prediction models translate to settings where data are collected primarily for operational purposes with irregular follow-up, limited predictors, and uncertain data quality has received little attention. This question matters most in resource-limited healthcare systems, where routine electronic health records (EHR) data are often the most feasible source for prediction model development.

In this study, we develop and internally validate a dynamic prediction model for VF using routine EHR data from PWH receiving ART in the Western Cape, South Africa.

## 2. Methods

### 2.1 Study Design and Population

This retrospective cohort study used data from the National Health Laboratory Service (NHLS) in the Western Cape, SA including all PWH attending public sector HIV care between January 1, 2008, and September 30, 2018. Data were obtained from the NHLS National Priority Programme HIV viral load unit and comprised individual identifiers, visit dates, demographic characteristics (date of birth and sex at birth), and HIV viral load results.

Records were linked (see below) and were later excluded in the analysis if individuals were younger than 16 years at first HIV viral load test, attended non-public healthcare facilities, or had fewer than three recorded viral load measurements in the follow up period (**Figure 1**). After exclusions, individuals were randomly allocated to a development cohort (75%) and a validation cohort (25%).

**Figure 1:**
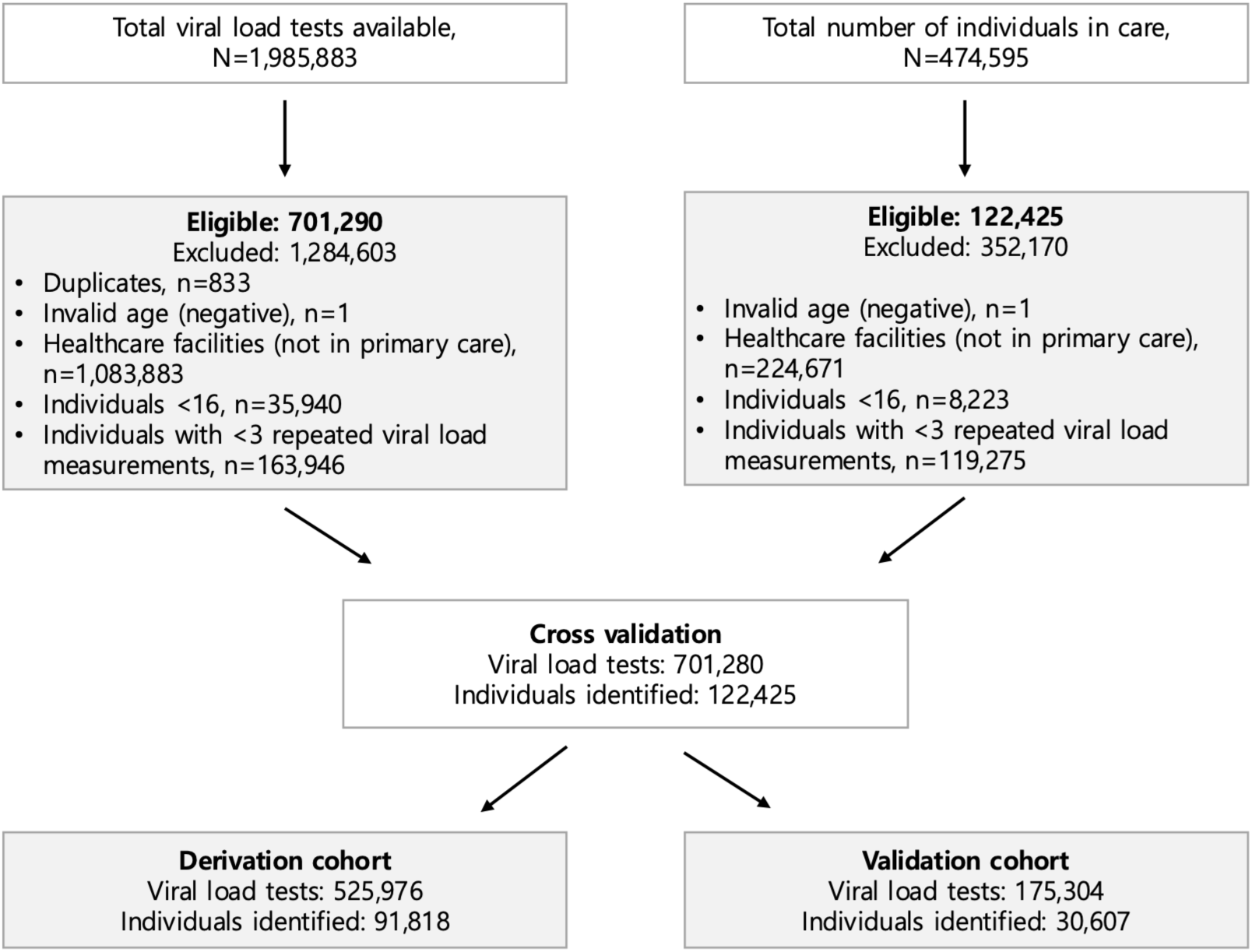
Flow diagram of individuals in the retrospective cohort study from the Western Cape, South Africa after data linkage.

### 2.2 Data and Record Linkage

Unlinked HIV viral load test results were linked using the record linkage procedure reported in Mukonda et al. [26], which carried out a multistage linkage using identifiers such as name/partial name, date of birth, and hospital identifier. Sensitivity analyses confirmed good linkage performance. [26] Individuals in this study were only identified by unique anonymous study identifier and had complete records.

### 2.3 Clinical Outcome and Longitudinal Biomarker

The clinical outcome of interest was time to first VF, defined as two consecutive HIV viral load (HIVVL) measurements ≥1000 copies/mL two-six months apart, following at least six months on ART. This definition is consistent with 2016 World Health Organization (WHO) and 2015 SA’s ART guidelines [27, 28], which were current at the time of data collection. Baseline was defined as the first recorded viral load measurement date. Total follow-up was calculated in weeks to either VF or censoring; individuals were censored at their last measurement if lost to follow-up or VF-free at study end. The longitudinal outcome was repeated HIVVL measurement. Given the right-skewed distribution of HIVVL values, all measurements were log base 10 transformed for all modelling but have been back transformed for reporting in text.

Viral load results reported below standard assay thresholds were handled following the approach of Hardie et al.[29] where results below 10 copies/mL were assigned a value of 9 copies/mL, results below 20 copies/mL were assigned 19 copies/mL, and results below 50 copies/mL were assigned 49 copies/mL. Where a quantified result was available, the recorded value was used directly.

### 2.4 Joint Model Specification

The association between the longitudinal log HIVVL trajectory and time to first VF was modelled using a joint model comprising two linked sub-models.

#### Longitudinal sub-model

Two linear mixed-effects (LME) structures were compared: random intercepts only, and random intercepts with linear random slopes. Both included a nonlinear effect of follow-up time through B-splines (internal knots at the 5th [week 0] and 95th [week 317] percentiles), and baseline age group (≤34, 35-44, ≥45 years) and sex at birth as fixed-effects. The model takes the form:

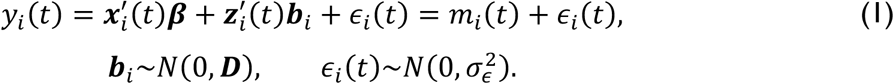

where *y_i_*(*t*) is the observed log HIVVL for individual *i*, *i* = 1, ⋯, *n*, at time *t*; *m_i_*(*t*) is the underlying true trajectory; ***x****_i_*(*t*) and ***z****_i_*(*t*) are fixed and random-effects design vectors for parameters ***β*** and ***b****_i_*; ***b****_i_* with unstructured variance-covariance matrix ***D***, respectively; and *ε_i_*(*t*) are residuals independent of ***b****_i_*. Model fit was compared using the Bayesian Information Criterion (BIC), with lower BIC value indicating better fit [30].

#### Event sub-model

The hazard of first VF was specified as:

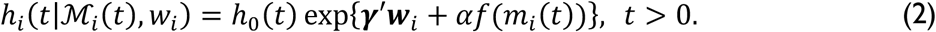

where ℎ_0_(*t*) is a B-spline baseline hazard; ***w****_i_* is a vector of baseline covariates with coefficients ***γ***; ℳ*_i_*(*t*) is a complete history of the true log HIVVL up to time *t*; and *α* quantifies the association between the log HIVVL trajectory and VF hazard. Three association structures were compared: (i) current value, *f*(*m_i_*(*t*)) = *m_i_*(*t*), which links the hazard at time *t* directly to the current underlying log HIVVL trajectory; (ii) lagged effects, *f*(*m_i_*(*t*)) = *m_i_*(max(*t* − *c*, 0)), which links the hazard to the log HIVVL trajectory six months prior, where *c* is a six-month lag; and iii) current value plus slope, 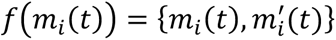, which additionally incorporates the instantaneous rate of change in the log HIVVL trajectory at time *t*, where 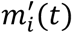 is the first derivative of the true trajectory.

All joint models were estimated using Bayesian Markov Chain Monte Carlo (MCMC) simulation [21, 31, 32]. Non-informative priors, three chains, and 2000 post-burn-in samples per parameter were used. Model fit was compared using the deviance information criterion (DIC) [32, 33], the Watanabe-Akaike information criterion (WAIC) [34], and the log-pseudo-marginal-likelihood (LPML) [35], with lower DIC and WAIC and higher LPML values indicating better fit.

### 2.5 Dynamic Prediction

The conditional probability that individual *j* remains VF-free beyond future time *u*, given survival to time *t* and accumulated viral load history *y*_j_(*t*) = {*y*_j_(*s*); 0 ≤ *s* ≤ *t*}, is:

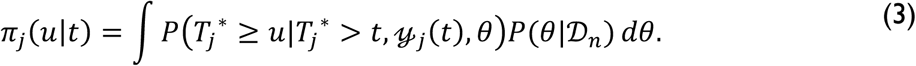

where 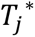 is the true event time; *θ* is the full parameter vector and *D_n_* is the development data. This integral is approximated through MCMC draws from the posterior of *θ*, yielding an estimated VF-free probability *π̂_j_*(*u*|*t*), updated each time a new viral load measurement becomes available [16, 19, 21, 36, 37].

### 2.6 Predictive Performance and Illustration

The data was split into a development cohort and a holdout validation cohort. Predictive performance was evaluated in the holdout validation cohort among individuals VF-free at six months, using data accumulated over that period to predict VF at 12 and 24 months. Discrimination was assessed using time-dependent area under the receiver operating characteristic curve (AUC) [16, 38], with values closer to 1 indicating better ability to distinguish individuals who will and will not experience VF; calibration through Brier scores [39, 40], with values closer to 0 indicating lower prediction error and better agreement between predicted and observed time-to-VF. Conditional VF probabilities were estimated at each follow-up, updated with each new measurement, and plotted with 95% credible intervals. All analyses were conducted in R software [41], using the nlme [42], survival [43] and JMbayes2 [44] packages. The study followed the Strengthening the Reporting of Observational studies in Epidemiology (STROBE) guideline[45] (**Supplementary Table S1**).

### 2.7 Ethics considerations

Ethical approval was granted by the Human Research Ethics Committee (HREC) of the University of Cape Town (Protocol No. HREC 436-2020). Informed consent was waived given the retrospective use of de-identified data.

## 3. Results

### 3.1 Cohort Characteristics

The development cohort had 91,818 individuals and 525,976 observations, while the validation cohort consisted of 30,607 individuals and 175,304 observations (**Figure 1**). Overall, females comprised 70% of included individuals, and 55% were younger than 35 years at enrolment (**Table 1**). At baseline, 71% had viral load below 50 copies/mL and 14% had ≥1000 copies/mL.

**Table 1:** Baseline characteristics of the retrospective cohort from the Western Cape, South Africa.

| Characteristic | N = 122,425<sup>i</sup> |
| --- | --- |
| Baseline HIV viral load, n (%) |  |
| <50 copies/mL | 87,050 (71.1) |
| ≥50 & <400 copies/mL | 15,596 (12.7) |
| ≥400 & <1000 copies/mL | 2,645 (2.2) |
| ≥1000 copies/mL | 17,134 (14.0) |
| Baseline age group (years), n (%) |  |
| ≤34 | 67,389 (55) |
| 35-44 | 38,150 (31) |
| ≥45 | 16,886 (14) |
| Sex at birth, n (%) |  |
| Male | 37,185 (30) |
| Female | 85,240 (70) |
| Cohort enrolment, n (%) |  |
| 2008 | 9,061 (7.4) |
| 2009 | 7,352 (6.0) |
| 2010 | 7,079 (5.8) |
| 2011 | 11,777 (9.6) |
| 2012 | 14,675 (12) |
| 2013 | 16,243 (13) |
| 2014 | 15,426 (13) |
| 2015 | 19,325 (16) |
| 2016 | 16,352 (13) |
| 2017 | 4,913 (4.0) |
| 2018 | 222 (0.2) |

The median follow-up was 92 weeks (interquartile range (IQR): 29-192) and individuals contributed a median of five viral load measurements (IQR: 4-7) over the study period. By end of follow-up, 12,547 individuals (10.2%) had experienced their first VF and 109,878 (89.8%) were censored. No individual experienced VF on more than one occasion.

Among those who experienced VF, the youngest age group (≤34 years) accounted for the largest proportion (58%), and females made up the majority (68%) of VF cases (**Table 2**). Fewer than 5% of individuals in each annual enrolment cohort between 2008 and 2010 experienced VF.

**Table 2:** Characteristics of individuals at the time of virologic failure in the overall, development, validation cohorts from the Western Cape, South Africa.

| Characteristic | Overall cohort<br>N = 12,547<sup>1</sup> | Development cohort<br>N = 9,357<sup>1</sup> | Validation cohort<br>N = 3,190<sup>1</sup> |
| --- | --- | --- | --- |
| Baseline age group (years), n (%) |  |  |  |
| ≤34 | 7,308 (58) | 5,466 (58) | 1,842 (58) |
| 35-44 | 3,791 (30) | 2,802 (30) | 989 (31) |
| ≥45 | 1,448 (12) | 1,089 (12) | 359 (11) |
| Sex at birth, n (%) |  |  |  |
| Male | 4,009 (32) | 2,986 (32) | 1,023 (32) |
| Female | 8,538 (68) | 6,371 (68) | 2,167 (68) |
| Cohort enrolment, n (%) |  |  |  |
| 2008 | 205 (1.6) | 149 (1.6) | 56 (1.8) |
| 2009 | 478 (3.8) | 351 (3.8) | 127 (4.0) |
| 2010 | 523 (4.2) | 397 (4.2) | 126 (3.9) |
| 2011 | 1,282 (10) | 953 (10) | 329 (10) |
| 2012 | 1,249 (10.0) | 933 (10.0) | 316 (9.9) |
| 2013 | 1,584 (13) | 1,182 (13) | 402 (13) |
| 2014 | 1,448 (12) | 1,075 (11) | 373 (12) |
| 2015 | 1,373 (11) | 1,008 (11) | 365 (11) |
| 2016 | 1,747 (14) | 1,294 (14) | 453 (14) |
| 2017 | 1,702 (14) | 1,276 (14) | 426 (13) |
| 2018 | 956 (7.6) | 739 (7.9) | 217 (6.8) |

Individual trajectories for 20 randomly selected individuals, stratified by VF status (**Figure 2**) demonstrate the considerable between-individual variability and nonlinear time trends that motivate the use of flexible mixed-effects models with individual-specific random slopes. On average, individuals who experienced VF had higher and more variable viral loads over time, while those without VF maintained lower, more stable profiles (**Figure 2** and **Supplement Figure S1)**.

**Figure 2:**
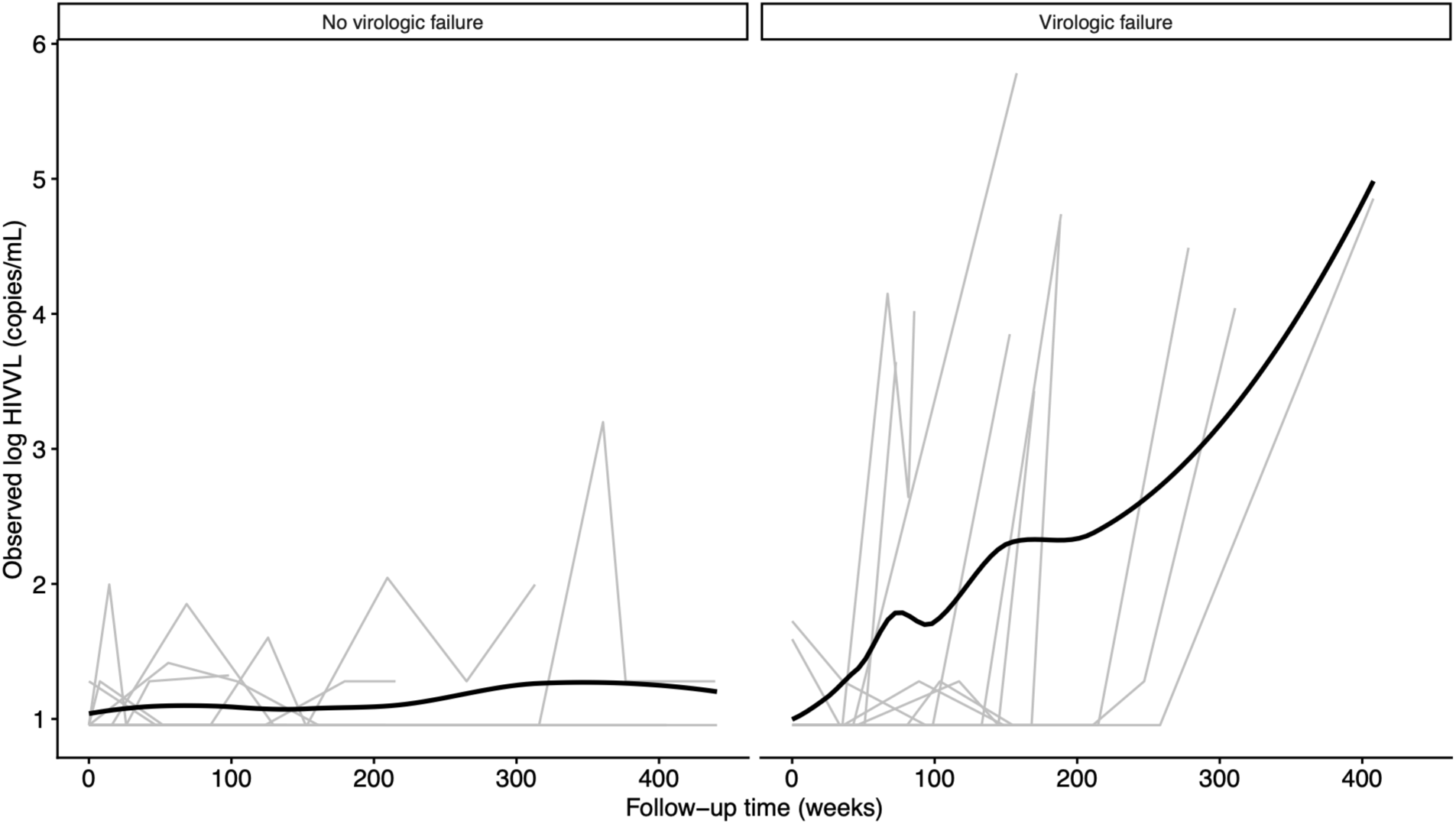
Exemplar longitudinal log HIVVL trajectories for 20 randomly selected individuals in the retrospective cohort from the Western Cape, South Africa, by virologic failure status. Abbreviations: HIVVL, HIV viral load.

The Kaplan-Meier (KM) estimate (**Supplement Figure S2**) estimated high VF-free probability throughout follow-up: 0.949 (95% confidence interval, CI: 0.948, 0.950) at 26 weeks (six months), 0.937 (95% CI: 0.936, 0.938) at 52 weeks (one year), 0.920 (95% CI: 0.918, 0.921) at 104 weeks (two years), 0.880 (95% CI: 0.878, 0.882) at 260 weeks (five years), and 0.830 (95% CI: 0.820, 0.840) at the end of follow-up (11 years).

### 3.2 Model Selection

The LME model with random intercepts and linear random slopes fitted considerably better than the random intercepts-only model (**Table 3**) and was carried forward. Among the joint models, the current value parameterization was preferred (**Table 3, Supplementary Figure S3**), and all subsequent analyses used this parameterization.

**Table 3:** Comparison of linear mixed-effects models and joint model association structures.

| Model | BIC | DIC | WAIC | LPML |
| --- | --- | --- | --- | --- |
| **Linear mixed-effects model** |  |  |  |  |
| Random intercepts only | 987,110.3 | - | - | - |
| Random intercepts and linear slopes | 982,999.1 | - | - | - |
| **Joint model** |  |  |  |  |
| Current value | - | 1,075,804.3 | 1,075,833.2 | -537,916.6 |
| Lagged effects | - | 1,243,524.7 | 1,085,634.9 | -542,823.3 |
| Current value plus Slope | - | 1,075,222.1 | 1,075,297.9 | -537,649.0 |
| Abbreviations: BIC, Bayesian information criterion; DIC, deviance information criterion; WAIC, Watanabe-Akaike information criterion; LPML, log-pseudo-marginal-likelihood value |  |  |  |  |

In the longitudinal sub-model, males had higher log HIVVL on average (beta: 0.107; 95% credible intervals, CrI: 0.095, 0.118) than females, and younger individuals (≤34 years) had consistently higher viral loads than older groups (**Table 4, Supplement Table S1**). The B-spline terms captured a nonlinear decline in log HIVVL over time, most prominently in the second spline coefficient (beta: −0.380; 95% CrI: −0.395, −0.365).

**Table 4:** Parameter estimates and 95% credible intervals from current value-parameterized joint model.

|  | Posterior<br>Mean | Standard<br>Error | 95% Credible Interval |  |
| --- | --- | --- | --- | --- |
|  |  |  | 2.5% | 97.5% |
| Time-to-event process |  |  |  |  |
| Baseline age group (years) |  |  |  |  |
| ≤34 | - | - | - | - |
| 35-44 | -0.017 | 0.00575 | -0.168 | 0.121 |
| ≥45 | -0.147 | 0.00685 | -0.288 | 0.002 |
| Sex at birth |  |  |  |  |
| Female | - | - | - | - |
| Male | -0.602 | 0.00397 | -0.700 | -0.513 |
| log HIV viral load (copies/mL) | 6.164 | 0.017 | 6.019 | 6.270 |
| Longitudinal process |  |  |  |  |
| Intercept | 1.681 | 0.000153 | 1.673 | 1.690 |
| Follow-up time (weeks): Spline 1 | -0.007 | 0.000280 | -0.020 | 0.005 |
| Follow-up time (weeks): Spline 2 | -0.380 | 0.000452 | -0.395 | -0.365 |
| Follow-up time (weeks): Spline 3 | -0.041 | 0.000688 | -0.057 | -0.025 |
| Baseline age group (years) |  |  |  |  |
| ≤34 | - | - | - | - |
| 35-44 | -0.053 | 0.000162 | -0.064 | -0.042 |
| ≥45 | -0.058 | 0.000204 | -0.074 | -0.043 |
| Sex at birth |  |  |  |  |
| Female | - | - | - | - |
| Male | 0.107 | 0.000144 | 0.095 | 0.118 |

In the event sub-model, each log increase in log HIVVL was associated with a 6.16-fold increase in the log hazard of VF (95% CrI: 6.02, 6.27), after adjusting for age group and sex at birth. Males had lower hazard of VF than females (log hazard ratio: −0.602; 95% CrI: −0.70, −0.513).

### 3.3 Predictive Performance and Dynamic Prediction Illustration

The final model was applied to the hold out validation cohort and evaluated. Discrimination was moderate (AUC: 0.69 at 12 months, 0.73 at 24 months). Calibration was good (Brier scores: 0.033 and 0.051 at 12 and 24 months), indicating that predicted VF probabilities closely matched observed VF rates at both time horizons (**Table 5**).

**Table 5:** Current value-parameterised joint model’s predictive performance at selected time windows in the validation cohort.

| Time window | AUC | Brier score |
| --- | --- | --- |
| 12-month | 0.69 | 0.033 |
| 24-month | 0.73 | 0.051 |
| *Abbreviations: AUC, Area under the curve.* |  |  |

A typical example of the model performing in an individual from the validation cohort is shown in **Figure 3** with four ‘snapshots’ at different time points showing the updated estimate of probability of VF. Credible intervals were relatively wide, reflecting imprecision in individual trajectory estimates.

**Figure 3:**
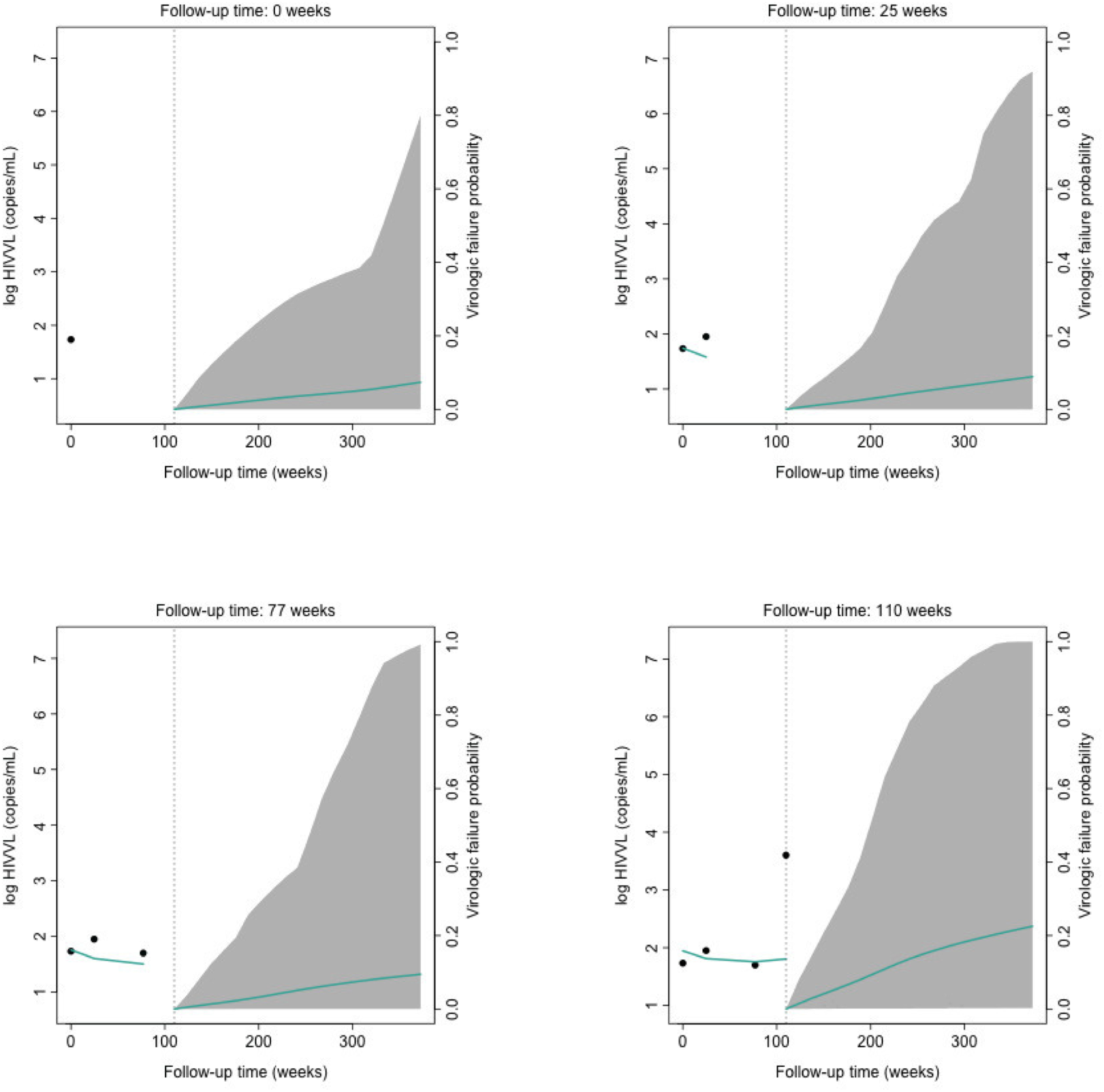
Dynamic virologic failure (VF) predictions: VF occurred at 110 weeks. Note: The x-axis shows follow-up time, with vertical dotted line marking the latest measurement. The left y-axis represents available log HIVVL up to the latest follow-up. Points show observed values, and solid green line represents the fitted longitudinal trajectory. The right y-axis shows estimated VF probabilities (solid green lines) and the corresponding 95% credible interval (shaded area). Abbreviations: HIVVL, HIV viral load.

## 4. Discussion

In this study, virologic failure affected 10.2% of individuals, within the 8-22% range reported from South Africa’s national treatment programme [15, 46, 47] and is consistent with comparable resource-limited settings [48, 49]. The low rate in the Western Cape likely reflects its strong healthcare infrastructure [50], but VF’s consequences, including regimen switching, increased costs, and deteriorating health outcomes, remain clinically and economically significant even at lower incidence [51, 52]. The absence of dynamic prediction tools for VF underscores the practical value of this work.

Three association structures were evaluated. The current value plus slope parameterization had lowest DIC and WAIC. However, model fit alone did not determine the structure, as plausibility depends on data characteristics. With sparse and irregular follow-up, individual slope estimates are likely imprecise, and reflecting measurement noise, risking overfitting. The current value parameterization was retained as the most biologically and statistically sensible choice. It is also the most clinically coherent structure for HIV viral load under ART. Viral load fluctuations in ART-treated individuals typically reflect adherence lapses close to measurements [3], making the current trajectory a more informative signal of VF than its value six months prior or its rate of change estimated from sparse data. The finding of a 6.2-fold increase in VF log hazard per log increase in log HIVVL is consistent with evidence from SSA showing that viral load changes predict adverse treatment outcomes [53]. Zakaria et al. [53] used a lagged parameterization, suggesting that the optimal association structure may vary by context. Our findings identified the current value as the stronger signal. Regardless of parameterization, both studies reinforce a shared conclusion: longitudinal viral load data substantially improve VF risk characterization over baseline-only approaches [14, 15], providing empirical support for dynamic prediction modelling in this application.

The discrepancy between the longitudinal and event sub-models regarding sex at birth warrants careful interpretation. In the longitudinal sub-model, males had higher average log HIVVL than females, while in the event sub-model, males had a lower hazard of VF after adjusting for the current log HIVVL. This is not a statistical contradiction; the two sub-models answer distinct questions. However, it reflects a well-recognized phenomenon in HIV care in SSA: men disengage from services at higher rates than women [54–56]. This suggests that those in care may have a lower failure risk than the broader male HIV-positive population. In this study, females made up the majority (68%) of VF cases, so the lower male hazard likely reflects engagement in-care patterns rather than a protective biological effect and should be interpreted with caution.

The performance pattern of good calibration alongside moderate discrimination is attributable to three interrelated features of routine EHR data, not inherent limitations of joint modelling. First, key predictors like ART regimen, adherence behavior, and co-infection status are absent, which are known to strongly differentiate individual VF risk [15, 51, 57, 58]. Routine EHR data collected for clinical documentation rarely capture detailed covariate information found in research cohorts [59, 60], limiting discriminative ability. Second, sparse and irregular measurement intervals make it difficult to reliably estimate individual-specific viral load trajectories from the LME, a prerequisite for accurate dynamic predictions [19]. The wide credible intervals observed for the illustrated individual reflect this imprecision. Third, the chronic nature of HIV and recommended follow-up of six to twelve months under South African guidelines [27, 28] at the time create measurement gaps that are often too wide to capture the interim viral load dynamics preceding VF, reducing the marginal information that longitudinal data contribute over a single baseline value. Together, these factors imply that the performance advantage commonly attributed to dynamic prediction models over other alternatives is not universal; it is conditional on the quality, frequency, and completeness of the data. This methodological insight advances the discussion on dynamic prediction modelling in resource-limited settings where routine EHR data are often the most available source, but where these structural challenges are most pronounced.

The few VF-specific prediction models that exist [14, 15], rely solely on baseline biomarker values, making direct comparisons of discrimination and calibration misleading. Broadly, dynamic prediction models evaluated in controlled settings benefit from systematic follow-up schedules, comprehensive covariate collection and high-quality data, which tend to produce favourable performance estimates than those achievable with EHR data. Therefore, the performance reported in this study should be viewed as a realistic estimate of what these models can achieve under real-world EHR data conditions, rather than as evidence of model inadequacy.

Despite moderate discrimination, the dynamic prediction illustration demonstrates genuine clinical utility. VF probabilities updated meaningfully at each visit, converging toward higher risk as VF approached. This capacity for continuous risk updating allows clinicians to reassess individual VF risk at each clinic contact rather than relying on a fixed initial estimate. In settings where viral load monitoring is the primary clinical tool and where treatment switches require careful justification, dynamically updated risk estimates can support timely decisions about enhanced adherence counselling or closer monitoring before VF is established [49]. Clinicians may not readily interpret posterior predicted probabilities, so a user-friendly implementation, for example, a web-based dynamic prediction calculator built using R Shiny [61], would be a necessary extension for practical deployment.

### Strengths and Limitations

The large sample size, over 120,000 individuals across a decade of routine care ensured sufficient statistical power and reflects the diversity of PWH in the Western Cape. All model predictors are routinely captured in EHR systems, making the model deployable without additional data collection infrastructure.

#### Several limitations exist

Missing clinical variables, including ART regimen, adherence, and co-infection status, limited discriminative performance and covariate coverage. Findings are specific to the Western Cape, with the lowest HIV prevalence (8.2%) [62] and strong healthcare infrastructure in South Africa [50]. Generalizability to higher-burden provinces with resource constraints is limited. External validation using data from other provinces or SSA healthcare settings is needed. The VF definition follows 2015 South African guidelines [27, 28], and findings should be interpreted cautiously under revised 2023 [63] and other guidelines. A sensitivity analysis using revised guidelines would be a useful extension of this work. The study period (2008-2018) predates the rollout of dolutegravir-based combination therapy in SA (2019 guidelines [8]). These regimens offer a higher genetic barrier to resistance and may produce different viral load trajectories and VF rates than the regimens in this cohort [8]. Applying the study’s model to more recent data would be a valuable extension. Finally, the joint modelling framework does not natively handle viral load values below the assay detection limit, which were present in this study. A two-part or robust joint model accommodating semicontinuous [64, 65] or left-censored data [66, 67] would be a worthwhile extension.

## 5. Conclusion

This study developed and internally validated the first dynamic prediction model for VF using routine HIV EHR data in South Africa. The joint modelling framework generated individual-specific, continuously updated risk estimates from real-world monitoring data, demonstrating that dynamic prediction is both feasible and clinically informative in this setting. While good calibration and meaningful dynamic predictions were achieved, moderate discrimination reflected the structural limitations of operationally collected data rather than a failure of the modelling approach.

Data quality, covariate coverage, and measurement frequency collectively determine what a dynamic prediction model can achieve and that these data characteristics can constrain performance even when the model is well-specified. This challenges the assumed superiority of dynamic prediction models and offers a clearer understanding of when these methods are most suitable. For the field of dynamic prediction modelling, evaluation in routine EHR settings is not merely an application exercise, it is essential for understanding the real-world boundaries of these methods. Future priorities include external validation in other resource-limited settings, incorporation of richer clinical predictors, intensification of recommended monitoring frequencies, and methodological extensions to accommodate left-censored or semicontinuous viral load values. With these developments, dynamic prediction models hold genuine promise to support personalized, timely clinical decision-making for PWH in resource-limited healthcare settings.

## Data Availability

The data are not publicly available due to privacy and ethical restrictions. The data are owned by the South African National Health Laboratory Service (NHLS) and include personal identifiers; requests for access should be directed to the NHLS.

## Acknowledgements

The authors thank the South African NHLS for providing the data used in this study. The content does not represent the official views of the NHLS. Part of this work was presented at the 62nd Annual Conference of the South African Statistical Association 2021, Stellenbosch, Western Cape, South Africa.

## Authors’ contributions

ML and FH conceptualized the study and developed the methodology and analysis plan. MH and EM contributed to data curation. FH conducted the formal statistical analyses. FH, ML, and FG contributed to interpreting the results of the formal analysis. FH drafted the original manuscript with support from ML. LM, EM, MH and FG provided both clinical and statistical expertise and contributed to manuscript revision. All authors reviewed and revised the manuscript, approved the final version for publication, and agree to be accountable for all aspects of the work, including the accuracy and integrity of the research.

## Availability of data and materials

The data are not publicly available due to privacy and ethical restrictions. The data are owned by the South African NHLS and include personal identifiers; requests for access should be directed to the NHLS.

## Reference

1. Kharsany, A.B. and Q.A. Karim, HIV infection and AIDS in sub-Saharan Africa: current status, challenges and opportunities. The open AIDS journal, 2016. 10: p. 34.

2. UNAIDS, D., Geneva: Joint United Nations Programme on HIV/AIDS. 2024.

3. WHO. Consolidated guidelines on HIV prevention, diagnosis, treatment and care for key populations–2016 update. 2016 17 Mar 2021]; Available from: https://www.who.int/publications/i/item/9789241511124.

4. WHO, Consolidated guidelines on the use of antiretroviral drugs for treating and preventing HIV infection: recommendations for a public health approach. 2016.

5. Saura-Lázaro, A., et al., Field performance and cost-effectiveness of a point-of-care triage test for HIV virological failure in Southern Africa. J Int AIDS Soc, 2023. 26(10): p. e26176.

6. Pham, M.D., et al., Viral load monitoring for people living with HIV in the era of test and treat: progress made and challenges ahead - a systematic review. BMC Public Health, 2022. 22(1): p. 1203.

7. Roberts, T., et al., Scale-up of Routine Viral Load Testing in Resource-Poor Settings: Current and Future Implementation Challenges. Clinical Infectious Diseases, 2016. 62(8): p. 1043–1048.

8. South African National Department of Health. 2019 ART Clinical Guidelines for the Management of HIV in Adults, Pregnancy, Adolescents, Children, Infants and Neonates. 2019 17 Mar 2021]; Available from: https://www.health.gov.za/hiv-and-aids/.

9. Hans, L., et al., HIV Viral Load Testing in the South African Public Health Setting in the Context of Evolving ART Guidelines and Advances in Technology, 2013-2022. Diagnostics (Basel), 2023. 13(17).

10. Han, X., et al., A nomogram for predicting paradoxical immune reconstitution inflammatory syndrome associated with cryptococcal meningitis among HIV-infected individuals in China. AIDS Res Ther, 2022. 19(1): p. 20.

11. Hou, X., et al., Development and validation of a prognostic nomogram for HIV/AIDS patients who underwent antiretroviral therapy: Data from a China population-based cohort. EBioMedicine, 2019. 48: p. 414–424.

12. Jiang, F., et al., Construction and validation of a prognostic nomogram for predicting the survival of HIV/AIDS adults who received antiretroviral therapy: a cohort between 2003 and 2019 in Nanjing. BMC Public Health, 2022. 22(1): p. 30.

13. Li, B., et al., A novel prediction model to evaluate the probability of CD4+/CD8+ cell ratio restoration in HIV-infected individuals. Aids, 2022. 36(6): p. 795–804.

14. Bewket, B., E.D. Abawa, and Z.A. Anteneh, Validation of a Risk Prediction Nomogram for Virologic Failure Among Patients on First-Line ART After ONE-J Program Implementation in Northwest Ethiopia. Journal of the International Association of Providers of AIDS Care (JIAPAC), 2026. 25: p. 23259582261428509.

15. Rohr, J.K., et al., Developing a predictive risk model for first-line antiretroviral therapy failure in South Africa. J Int AIDS Soc, 2016. 19(1): p. 20987.

16. Rizopoulos, D., Dynamic predictions and prospective accuracy in joint models for longitudinal and time-to-event data. Biometrics, 2011. 67(3): p. 819–29.

17. Rizopoulos, D., Joint models for longitudinal and time-to-event data: With applications in R. 2012: Chapman and Hall/CRC.

18. Putter, H. and H.C. Van Houwelingen, Landmarking 2.0: Bridging the gap between joint models and landmarking. Stat Med, 2022. 41(11): p. 1901–1917.

19. Rizopoulos, D., G. Molenberghs, and E.M. Lesaffre, Dynamic predictions with time-dependent covariates in survival analysis using joint modeling and landmarking. Biometrical Journal, 2017. 59(6): p. 1261–1276.

20. McHunu, N.N., et al., Optimizing personalized screening intervals for clinical biomarkers using extended joint models. J Appl Stat, 2026. 53(2): p. 171–202.

21. Proust-Lima, C. and J.M. Taylor, Development and validation of a dynamic prognostic tool for prostate cancer recurrence using repeated measures of posttreatment PSA: a joint modeling approach. Biostatistics, 2009. 10(3): p. 535–549.

22. Yu, M., J.M.G. Taylor, and H.M. Sandler, Individual prediction in prostate cancer studies using a joint longitudinal survival–cure model. Journal of the American Statistical Association, 2008. 103(481): p. 178–187.

23. Rueten-Budde, A.J., et al., Dynamic prediction of overall survival for patients with high-grade extremity soft tissue sarcoma. Surg Oncol, 2018. 27(4): p. 695–701.

24. Andrinopoulou, E., et al., Combined dynamic predictions using joint models of two longitudinal outcomes and competing risk data. Stat Methods Med Res, 2017. 26(4): p. 1787–1801.

25. Grand, M.K., et al., A joint model for dynamic prediction in uveitis. Stat Med, 2019. 38(10): p. 1802–1816.

26. Mukonda, E., et al., Mixed-method estimation of population-level HIV viral suppression rate in the Western Cape, South Africa. BMJ Global Health, 2020. 5(8): p. e002522.

27. WHO, Consolidated guidelines on the use of antiretroviral drugs for treating and preventing HIV infection: recommendations for a public health approach. - 2nd ed. 2016: World Health Organization.

28. South Africa National Department of Health, S., National consolidated guidelines for the prevention of mother-to-child transmission of HIV (PMTCT) and the management of HIV in children, adolescents and adults. 2015, NDoH Pretoria.

29. Hardie, D.R., et al., Field study to determine the reliability of HIV viral load results shows minimal impact of delayed testing in South Africa. 2024, 2024. 13(1).

30. Schwarz, G., Estimating the dimension of a model. The annals of statistics, 1978. 6(2): p. 461–464.

31. Rizopoulos, D., Dynamic predictions and prospective accuracy in joint models for longitudinal and time-to-event data. Biometrics, 2011. 67(3): p. 819–829.

32. Gilks, W.R., S. Richardson, and D. Spiegelhalter, Markov chain Monte Carlo in practice. 1995: CRC press.

33. Spiegelhalter, D.J., et al., Bayesian measures of model complexity and fit. Journal of the royal statistical society: Series b (statistical methodology), 2002. 64(4): p. 583–639.

34. Gelman, A., J. Hwang, and A. Vehtari, Understanding predictive information criteria for Bayesian models. Statistics and computing, 2014. 24(6): p. 997–1016.

35. Geisser, S. and W.F. Eddy, A predictive approach to model selection. Journal of the American Statistical Association, 1979. 74(365): p. 153–160.

36. Andrinopoulou, E., et al., Reflection on modern methods: dynamic prediction using joint models of longitudinal and time-to-event data. International Journal of Epidemiology, 2021. 50(5): p. 1731–1743.

37. Li, K. and S. Luo, Dynamic predictions in Bayesian functional joint models for longitudinal and time-to-event data: An application to Alzheimer’s disease. Statistical methods in medical research, 2019. 28(2): p. 327–342.

38. Heagerty, P. and Y. Zheng, Survival Model Predictive Accuracy and ROC Curves. Biometrics, 2005. 61(1): p. 92–105.

39. Gerds, T.A. and M. Schumacher, Consistent Estimation of the Expected Brier Score in General Survival Models with Right-Censored Event Times. Biometrical Journal, 2006. 48(6): p. 1029–1040.

40. Mogensen, U.B., H. Ishwaran, and T.A. Gerds, Evaluating Random Forests for Survival Analysis Using Prediction Error Curves. JOURNAL OF STATISTICAL SOFTWARE, 2012. 50(11): p. 1–23.

41. R Core Team, R: A Language and Environment for Statistical Computing. 2024, R Foundation for Statistical Computing: Vienna, Austria.

42. Pinheiro, J., et al., Package ‘nlme’. Linear and nonlinear mixed effects models, version, 2017. 3(1): p. 274.

43. Therneau, T.M. and T. Lumley, Package ‘survival’. R Top Doc, 2015. 128(10): p. 28–33.

44. Rizopoulos, D., G. Papageorgiou, and P. Miranda Afonso, JMbayes2: extended joint models for longitudinal and time-to-event data. R package version 0.3-0, ed, 2022.

45. von Elm, E., et al., The Strengthening the Reporting of Observational Studies in Epidemiology (STROBE) statement: guidelines for reporting observational studies. J Clin Epidemiol, 2008. 61(4): p. 344–9.

46. Hermans, L.E., et al., Effect of HIV-1 low-level viraemia during antiretroviral therapy on treatment outcomes in WHO-guided South African treatment programmes: a multicentre cohort study. Lancet Infect Dis, 2018. 18(2): p. 188–197.

47. Fox, M.P., et al., Rates and predictors of failure of first-line antiretroviral therapy and switch to second-line ART in South Africa. J Acquir Immune Defic Syndr, 2012. 60(4): p. 428–37.

48. Boender, T.S., et al., Long-term Virological Outcomes of First-Line Antiretroviral Therapy for HIV-1 in Low- and Middle-Income Countries: A Systematic Review and Meta-analysis. Clin Infect Dis, 2015. 61(9): p. 1453–61.

49. Mesic, A., et al., Predictors of virological failure among people living with HIV receiving first line antiretroviral treatment in Myanmar: retrospective cohort analysis. AIDS Research and Therapy, 2021. 18(1): p. 16.

50. Statistics South Africa. Financial statistics of provincial government 2019/2020. 2021 17 June 2023]; Available from: https://www.statssa.gov.za/?p=14755.

51. SeyedAlinaghi, S., et al., Current ART, determinants for virologic failure and implications for HIV drug resistance: an umbrella review. AIDS Research and Therapy, 2023. 20(1): p. 74.

52. Agegnehu, C.D., M.W. Merid, and M.K. Yenit, Incidence and predictors of virological failure among adult HIV patients on first-line antiretroviral therapy in Amhara regional referral hospitals; Ethiopia: a retrospective follow-up study. BMC Infect Dis, 2020. 20(1): p. 460.

53. Zakaria, H.F., et al., Joint Modeling of Incidence of Unfavorable Outcomes and Change in Viral Load Over Time Among Adult HIV/AIDS Patients on Second-Line Anti-Retroviral Therapy, in Selected Public Hospitals of Addis Ababa, Ethiopia. HIV AIDS (Auckl), 2022. 14: p. 341–354.

54. Cohn, J., et al., Sex Differences in the Treatment of HIV. Current HIV/AIDS Reports, 2020. 17(4): p. 373–384.

55. Eyassu, M.A., T.M. Mothiba, and N.P. Mbambo-Kekana, Adherence to antiretroviral therapy among HIV and AIDS patients at the Kwa-Thema clinic in Gauteng Province, South Africa. 2016, 2016. 8(2).

56. Hlongwa, M., et al., Linkage to HIV care and early retention in HIV care among men in the ‘universal test-and-treat’ era in a high HIV-burdened district, KwaZulu-Natal, South Africa. BMC Health Serv Res, 2024. 24(1): p. 384.

57. Agegnehu, C.D., et al., Burden and Associated Factors of Virological Failure Among People Living with HIV in Sub-Saharan Africa: A Systematic Review and Meta-Analysis. AIDS Behav, 2022. 26(10): p. 3327–3336.

58. Cevik, M., C. Orkin, and P.E. Sax, Emergent Resistance to Dolutegravir Among INSTI-Naïve Patients on First-line or Second-line Antiretroviral Therapy: A Review of Published Cases. Open Forum Infectious Diseases, 2020. 7(6).

59. Wright, A., F.L. Maloney, and J.C. Feblowitz, Clinician attitudes toward and use of electronic problem lists: a thematic analysis. BMC Med Inform Decis Mak, 2011. 11: p. 36.

60. Savitz, S.T., et al., How much can we trust electronic health record data? Healthcare, 2020. 8(3): p. 100444.

61. Wickham, H., Mastering shiny. 2021: ” O’Reilly Media, Inc.“.

62. Human Sciences Research Council. The Sixth South African National Hiv Prevalence, Incidence, Behaviour And Communication Survey, 2022. 2023 17 June 2023]; Available from: https://hsrc.ac.za/special-projects/sabssm-survey-series/sabssmvi-media-pack-november-2023/.

63. South African National Department of Health, ART Clinical Guidelines for the Management of HIV in Adults, Pregnancy and Breastfeeding, Adolescents, Children, Infants and Neonates, 2023. National ART Clinical Guideline, 2023.

64. Liu, L., Joint modeling longitudinal semi-continuous data and survival, with application to longitudinal medical cost data. Statistics in medicine, 2009. 28(6): p. 972–986.

65. Dagne, G.A., Joint two-part Tobit models for longitudinal and time-to-event data. Stat Med, 2017. 36(26): p. 4214–4229.

66. Król, A., et al., Joint model for left-censored longitudinal data, recurrent events and terminal event: Predictive abilities of tumor burden for cancer evolution with application to the FFCD 2000–05 trial. Biometrics, 2016. 72(3): p. 907–916.

67. Yu, T., et al., Robust joint modelling of left-censored longitudinal data and survival data with application to HIV vaccine studies. Ann Appl Stat, 2023. 17(2): p. 1017–1037.

