## Supplementary Material for "Dynamic Prediction to Predict Virologic Failure in Antiretroviral Treated People Living with HIV: Cohort Analysis of Routine Electronic Health Records Data in the Western Cape, South Africa"

[1]Division of Epidemiology and Biostatistics, School of Public Health, University of Cape Town, Cape Town, South Africa, [2]Department of Statistical Sciences, Faculty of Science, University of Cape Town, South Africa, [3]Division of Medical Virology, National Health Laboratory Service, University of Cape Town and Groote Schuur Hospital, Cape Town, South Africa, [4]National Heart and Lung Institute, Imperial College London, London, UK

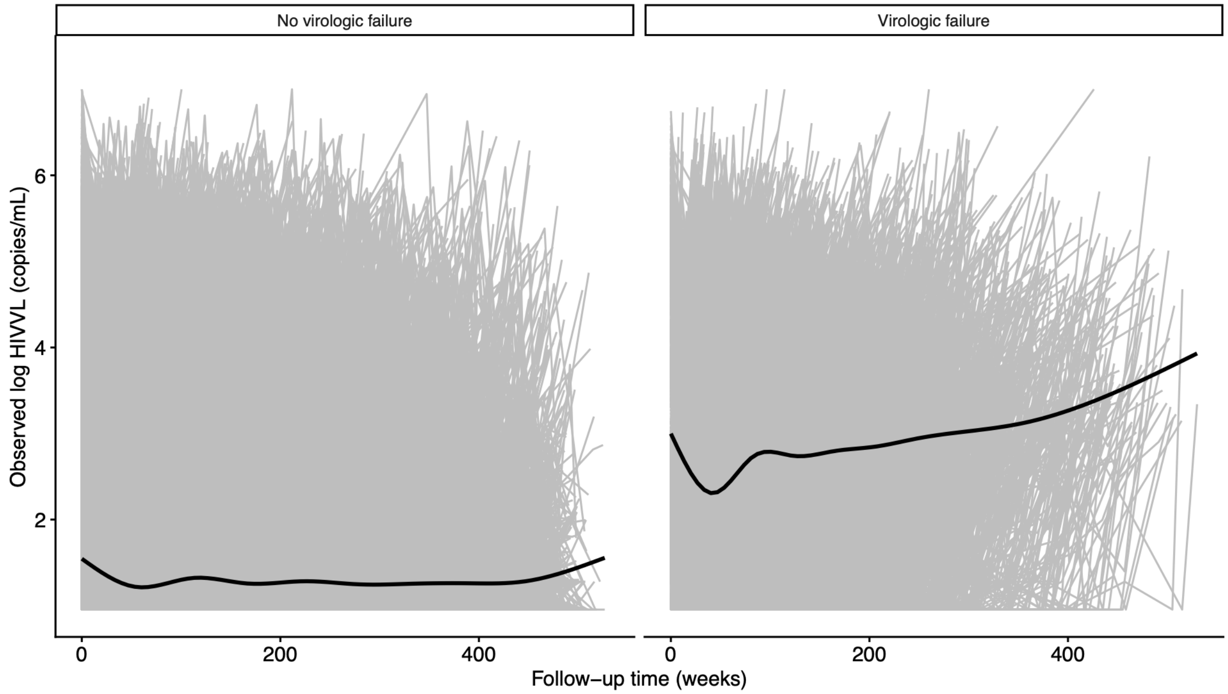


**Additional file 1. Figure S1**: Longitudinal log HIVVL trajectories over follow-up in the retrospective cohort from the Western Cape, South Africa, by virologic failure status. Abbreviations: HIVVL, HIV viral load.

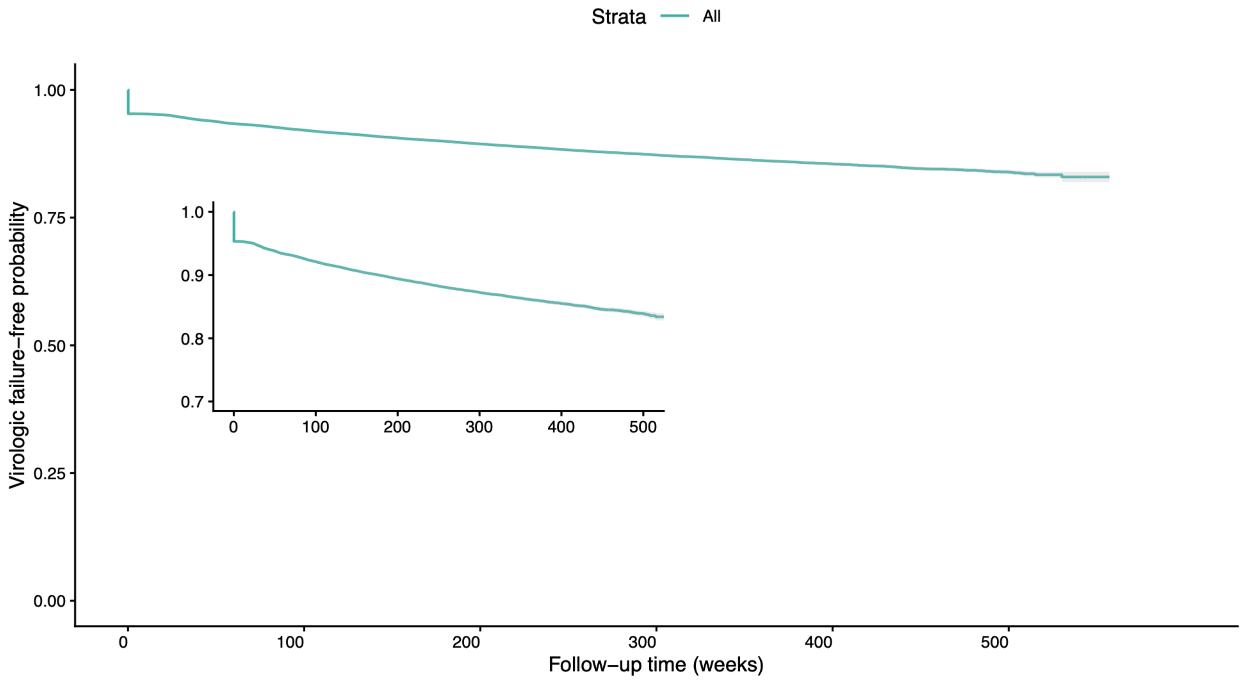


**Additional file 2. Figure S2**: Kaplan-Meier estimate of virologic failure-free probability in the retrospective cohort from the Western Cape, South Africa.

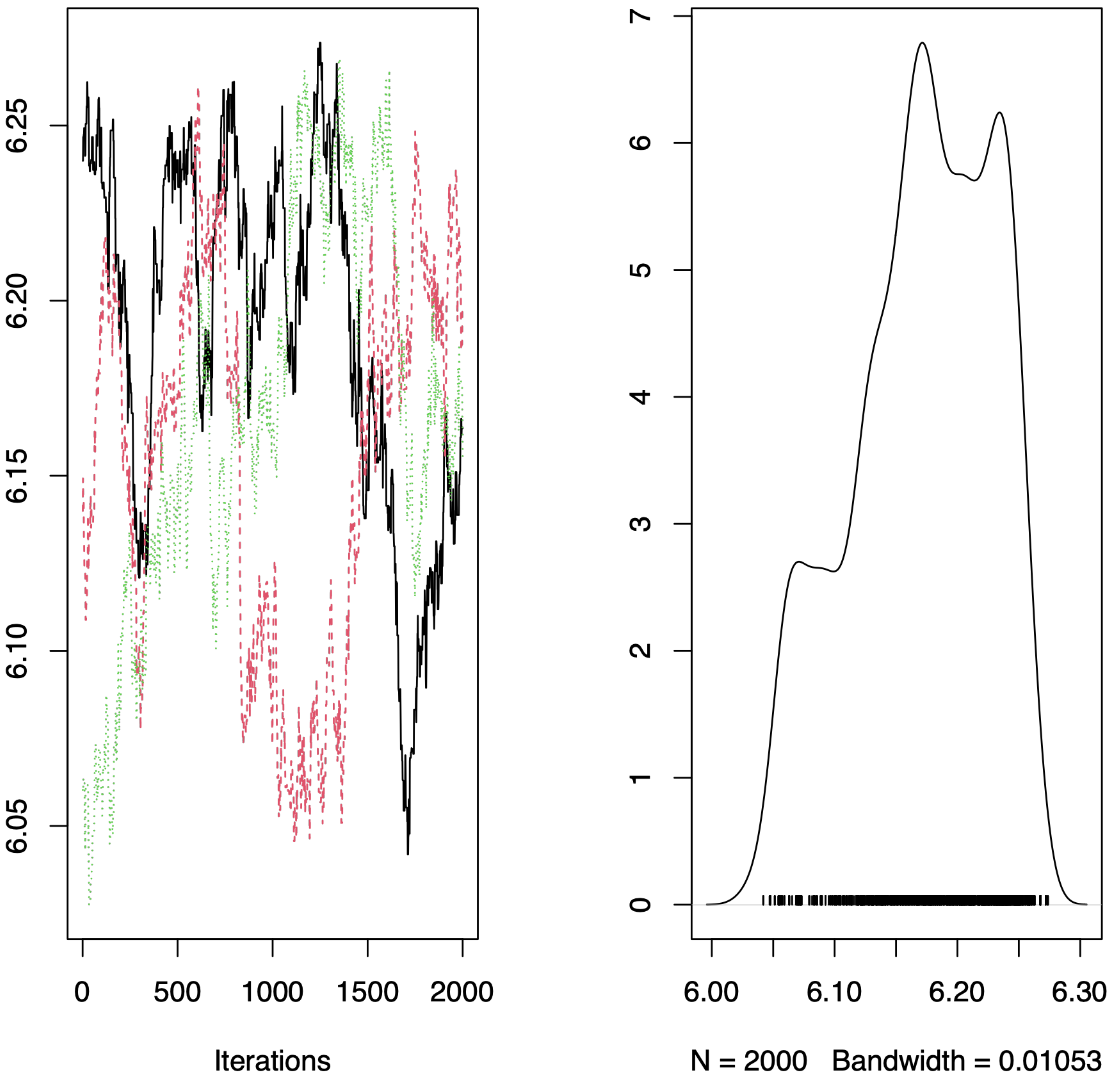


**Additional file 3 Figure S3**: Trace (left panel) and kernel density (right panel) plots for the current value-parameterized joint model.

### **Additional file 4. Table S1**: The STROBE Statement.

| | Item No | Recommendation | Reported |
|---|---|---|---|
| **Title and abstract** | 1 | (*a*) Indicate the study's design with a commonly used term in the title or the abstract | ✓ |
| | | (*b*) Provide in the abstract an informative and balanced summary of what was done and what was found | ✓ |
| **Introduction** | | | |
| Background/rationale | 2 | Explain the scientific background and rationale for the investigation being reported | ✓ |
| Objectives | 3 | State specific objectives, including any prespecified hypotheses | ✓ |
| **Methods** | | | |
| Study design | 4 | Present key elements of study design early in the paper | ✓ |
| Setting | 5 | Describe the setting, locations, and relevant dates, including periods of recruitment, exposure, follow-up, and data collection | ✓ |
| Participants | 6 | (*a*) *Cohort study*—Give the eligibility criteria, and the sources and methods of selection of participants. Describe methods of follow-up<br>*Case-control study*—Give the eligibility criteria, and the sources and methods of case ascertainment and control selection. Give the rationale for the choice of cases and controls<br>*Cross-sectional study*—Give the eligibility criteria, and the sources and methods of selection of participants | ✓ |
| | | (*b*) *Cohort study*—For matched studies, give matching criteria and number of exposed and unexposed<br>*Case-control study*—For matched studies, give matching criteria and the number of controls per case | |
| Variables | 7 | Clearly define all outcomes, exposures, predictors, potential confounders, and effect modifiers. Give diagnostic criteria, if applicable | ✓ |
| Data sources/ measurement | 8* | For each variable of interest, give sources of data and details of methods of assessment (measurement). Describe comparability of assessment methods if there is more than one group | ✓ |
| Bias | 9 | Describe any efforts to address potential sources of bias | |
| Study size | 10 | Explain how the study size was arrived at | ✓ |
| Quantitative variables | 11 | Explain how quantitative variables were handled in the analyses. If applicable, describe which groupings were chosen and why | ✓ |
| Statistical methods | 12 | (*a*) Describe all statistical methods, including those used to control for confounding | ✓ |
| | | (*b*) Describe any methods used to examine subgroups and interactions | |
| | | (*c*) Explain how missing data were addressed | ✓ |
| | | (*d*) *Cohort study*—If applicable, explain how loss to follow-up was addressed<br>*Case-control study*—If applicable, explain how matching of cases and controls was addressed<br>*Cross-sectional study*—If applicable, describe analytical methods taking account of sampling strategy | ✓ |
| | | (*e*) Describe any sensitivity analyses | |

Continued on next page

| Results | | | Reported |
|---|---|---|---|
| Participants | 13* | (a) Report numbers of individuals at each stage of study—eg numbers potentially eligible, examined for eligibility, confirmed eligible, included in the study, completing follow-up, and analysed | ✓ |
| | | (b) Give reasons for non-participation at each stage | |
| | | (c) Consider use of a flow diagram | ✓ |
| Descriptive data | 14* | (a) Give characteristics of study participants (eg demographic, clinical, social) and information on exposures and potential confounders | ✓ |
| | | (b) Indicate number of participants with missing data for each variable of interest | |
| | | (c) *Cohort study*—Summarise follow-up time (eg, average and total amount) | ✓ |
| Outcome data | 15* | *Cohort study*—Report numbers of outcome events or summary measures over time | ✓ |
| | | *Case-control study*—Report numbers in each exposure category, or summary measures of exposure | |
| | | *Cross-sectional study*—Report numbers of outcome events or summary measures | |
| Main results | 16 | (*a*) Give unadjusted estimates and, if applicable, confounder-adjusted estimates and their precision (eg, 95% confidence interval). Make clear which confounders were adjusted for and why they were included | ✓ |
| | | (*b*) Report category boundaries when continuous variables were categorized | |
| | | (*c*) If relevant, consider translating estimates of relative risk into absolute risk for a meaningful time period | |
| Other analyses | 17 | Report other analyses done—eg analyses of subgroups and interactions, and sensitivity analyses | ✓ |
| **Discussion** | | | |
| Key results | 18 | Summarise key results with reference to study objectives | ✓ |
| Limitations | 19 | Discuss limitations of the study, taking into account sources of potential bias or imprecision. Discuss both direction and magnitude of any potential bias | ✓ |
| Interpretation | 20 | Give a cautious overall interpretation of results considering objectives, limitations, multiplicity of analyses, results from similar studies, and other relevant evidence | ✓ |
| Generalisability | 21 | Discuss the generalisability (external validity) of the study results | ✓ |
| **Other information** | | | |
| Funding | 22 | Give the source of funding and the role of the funders for the present study and, if applicable, for the original study on which the present article is based | ✓ |

*Give information separately for cases and controls in case-control studies and, if applicable, for exposed and unexposed groups in cohort and cross-sectional studies.
